# Cohort-based surveillance of SARS-CoV2 transmission mirrors infection rates at the population level: a one-year longitudinal study

**DOI:** 10.1101/2021.05.10.21256966

**Authors:** Christine Klein, Max Borsche, Alexander Balck, Bandik Föh, Johann Rahmöller, Elke Peters, Jan Knickmann, Miranda Lane, Eva-Juliane Vollstedt, Susanne A. Elsner, Nadja Käding, Susanne Hauswaldt, Tanja Lange, Jennifer E. Hundt, Selina Lehrian, Julia Giese, Alexander Mischnik, Stefan Niemann, Florian Maurer, Susanne Homolka, Laura Paulowski, Jan Kramer, Christoph Twesten, Christian Sina, Gabriele Gillessen-Kaesbach, Hauke Busch, Marc Ehlers, Stefan Taube, Jan Rupp, Alexander Katalinic

**Affiliations:** Institute of Neurogenetics, University of Lübeck and University Hospital Schleswig-Holstein, Campus Lübeck, Lübeck, Germany; Department of Neurology, University of Lübeck, Lübeck, Germany; Institute of Nutritional Medicine, University of Lübeck, Lübeck, Germany; Medical Department I, University Hospital Schleswig-Holstein, Lübeck, Germany; Department of Anesthesiology and Intensive Care, University Medical Center Schleswig-Holstein, Lübeck, Germany; Institute of Social Medicine and Epidemiology, University of Lübeck, Lübeck, Germany; Institute of Virology and Cell Biology, University of Lübeck, Lübeck, Germany; Department of Infectious Diseases and Microbiology, University of Lübeck and University Hospital Schleswig-Holstein, Campus Lübeck, Lübeck, Germany; Department of Rheumatology & Clinical Immunology, University of Lübeck and University Hospital Schleswig-Holstein, Campus Lübeck, Lübeck, Germany; Lübeck Institute of Experimental Dermatology (LIED), University of Lübeck, Lübeck, Germany; Health Protection Authority, Lübeck, Germany; Research Center Borstel, Leibniz Lung Center, Borstel, Germany; German Center for Infection Research (DZIF), Partner site Hamburg-Lübeck-Borstel-Riems, Borstel, Germany; LADR Laboratory Group Dr. Kramer & Colleagues, Geesthacht, Germany; Perfood GmbH, Lübeck, Germany; University of Lübeck, Lübeck, Germany

**Keywords:** SARS-CoV-2, COVID-19, longitudinal study, surveillance, digital monitoring

## Abstract

**Background:** More than one year into the COVID-19 pandemic, important data gaps remain on longitudinal prevalence of SARS-CoV-2 infection at the population level and in defined risk groups, efficacy of specific lockdown measures, and on (cost-)effective surveillance.

**Methods:** The ELISA (Lüb**e**ck **L**ongitudinal **I**nvestigation of **SA**RS-CoV-2 Infection) study invited adult inhabitants (n=∼300,000) from the Lübeck area (Northern Germany) and enrolled 3051 participants (∼1%); 1929 population-matched and 1645 with high-exposure based on profession. The one-year study period (03/2020-02/2021) spanned massive influx of tourism in the summer, rise of infection rates in the fall/winter 2020/2021, and two lockdowns. Participants were screened seven times for SARS-CoV-2 infection using PCR and antibody testing and monitored with an app-based questionnaire (n=∼91,000).

**Results:** Cohort (56% female; mean age: 45.6 years) retention was 75%-98%; 89 persons (3.5%) were antibody- and/or PCR-positive. Seropositivity was almost 2-fold higher in men and increased risk detected in several high-exposure groups (highest for nurses, followed by police, army, firemen, and students). In May 2020, 92% of the infections were missed by PCR testing; by February 2021, only 29% remained undiagnosed. “Contact to COVID-19-affected” was the most relevant risk factor. Other factors, such as frequent use of public transportation, shopping, close contacts at work, and extensive tourism in the summer did not impact infection rates.

**Conclusions:** We i) provide a model for effective, regional surveillance; ii) identify infection risk factors informing public health measures; iii) demonstrate that easing of lockdown measures appears safe at times of low prevalence in the presence of continuous monitoring.

## Introduction

In their - still - timely Perspective entitled ‘Defining the epidemiology of COVID-19 – Studies needed’ published in March 2020, Lipsitch and colleagues called upon the scientific community to carry out studies providing the evidence required for controlling an epidemic.^1^ Over the past year, the epidemic has evolved into a pandemic with >170 million confirmed cases and >3.5 million deceased COVID-19 patients world-wide.^2^ Although research activities on all aspects of SARS-CoV-2 infection and COVID-19 are unprecedented in speed and scope, information currently remains scarce on i) prevalence and dynamics of SARS-CoV-2 infection and longitudinal antibody profiles at the population level and in defined risk groups; ii) efficacy of and compliance with different lockdown measures; and iii) (cost-)effective surveillance approaches that allow representative monitoring of the population including early detection of asymptomatic cases and new outbreaks especially in low-prevalence regions. Population-based studies, such as the one undertaken in the Icelandic population at the beginning of the pandemic, are typically cross-sectional or cover short time intervals.^3^ In the Italian municipality of Vo, initial testing of 86% of the population revealed a prevalence of SARS-CoV-2 infection of 2.6%, which dropped to 1.2% after a strict lockdown and revealed 42% of asymptomatic infections.^4^ However, the population’s behavior was not systematically monitored and long-term follow-up data are unavailable, as is the case for ongoing longitudinal studies for which only study protocols have been published to date.^5^ Of further note, effects of containment measures are rarely assessed directly but rather modeled^6,7,8,9,10^ and unlikely to be widely generalizable from one geographic and/or cultural setting to another.

We here report the results of a 1-year, longitudinal, prospective cohort study surveilling ∼3000 people (∼1% of the population living in our catchment area), which included ∼20,000 virus PCR and anti-SARS-CoV-2 spike protein (S1) IgG antibody tests and >90,000 detailed App-based data sets on symptoms, mobility, pandemic-related behavior, and quality of life.

## Methods

### Study oversight and design

The ELISA (Lüb**e**ck **L**ongitudinal **I**nvestigation of **SA**RS-CoV-2 Infection) study was approved by the ethics committee of the University of Lübeck (Az. 20-150). Participants were invited to take part in the study through announcements in the local press, posters and leaflets, and by advertisement on the official homepage of the City of Lübeck by its mayor. Detailed study information was provided on ELISA’s website (https://elisa-luebeck.de), by telephone counseling, and in newsletters. Within two weeks in April 2020, 7,303 adult inhabitants of the greater Lübeck area (approx. 300,000 inhabitants) downloaded the dedicated ELISA App. All 7303 participants were invited to fill out the study questionnaire, covering retrospectively also the period from March 2020; 3474 were selected for subsequent SARS-CoV-2 testing. Of these, 2145 individuals matched by age and sex distribution of the study region were randomly drawn to comprise a population-based group. An additional 1329 individuals were selected to enrich a potential high-exposure group based on profession requiring intense and/or frequent contact with other people, such as health care personnel (**Figure 1**). A total of 3051 persons (88%) came to the study center for initial screening (ELISA cohort). All cohort members were invited to the study center seven times between early May 2020 to February 2021 spanning the end of the first lockdown, massive influx of tourism in the summer, steep rise of infection rates in the fall/winter 2020/2021, and the second lockdown (**Figure 2.A**). All participants were tested for current or previous SARS-CoV-2 infection using nucleic-acid and anti-spike protein (extracellular S1 part) IgG antibody testing with support of a mobile app-based questionnaire, electronic scheduling and result reporting system.

**Figure 1.**
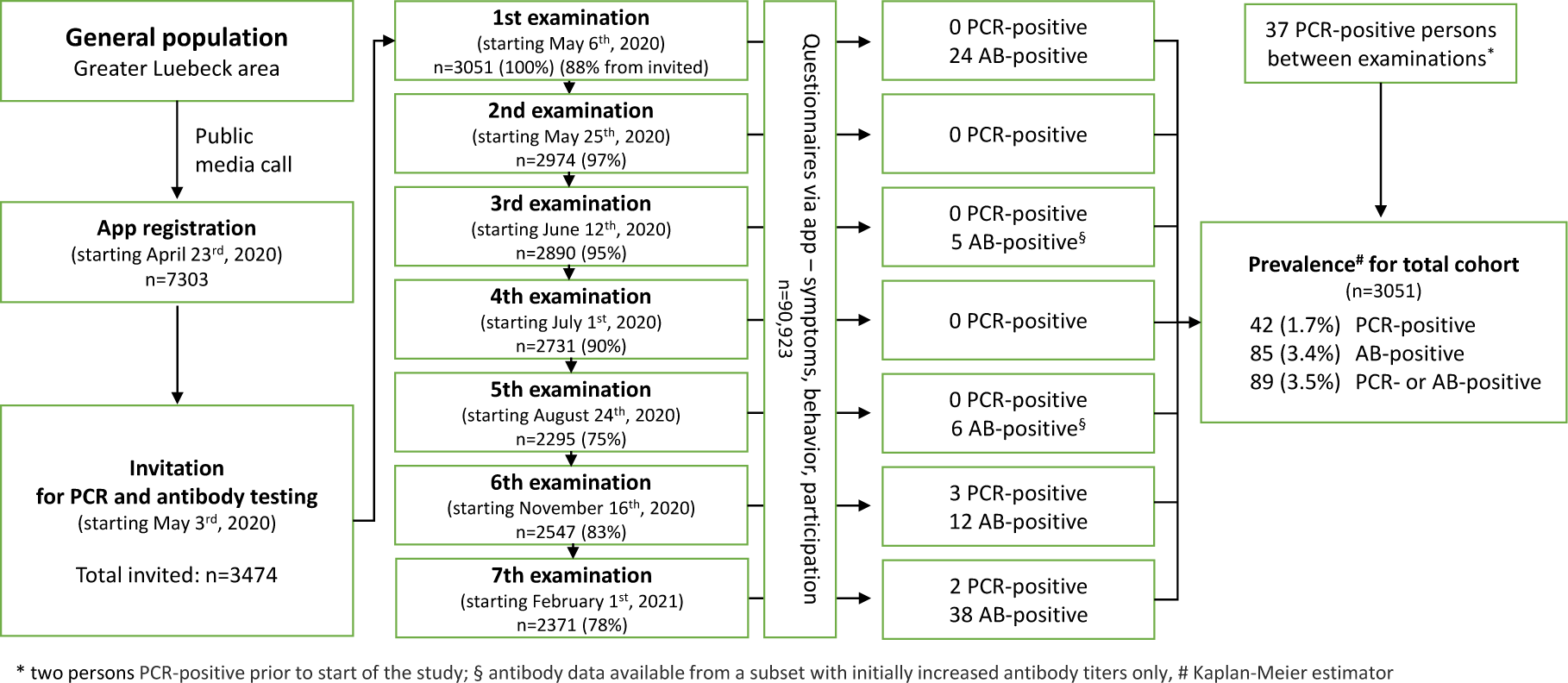
Study design and numbers of PCR- and anti-S1 IgG antibody (AB)-positive individuals. The ELISA cohort was prospectively followed from May 6^th^, 2020 – February 27^th^, 2021.

**Figure 2.**
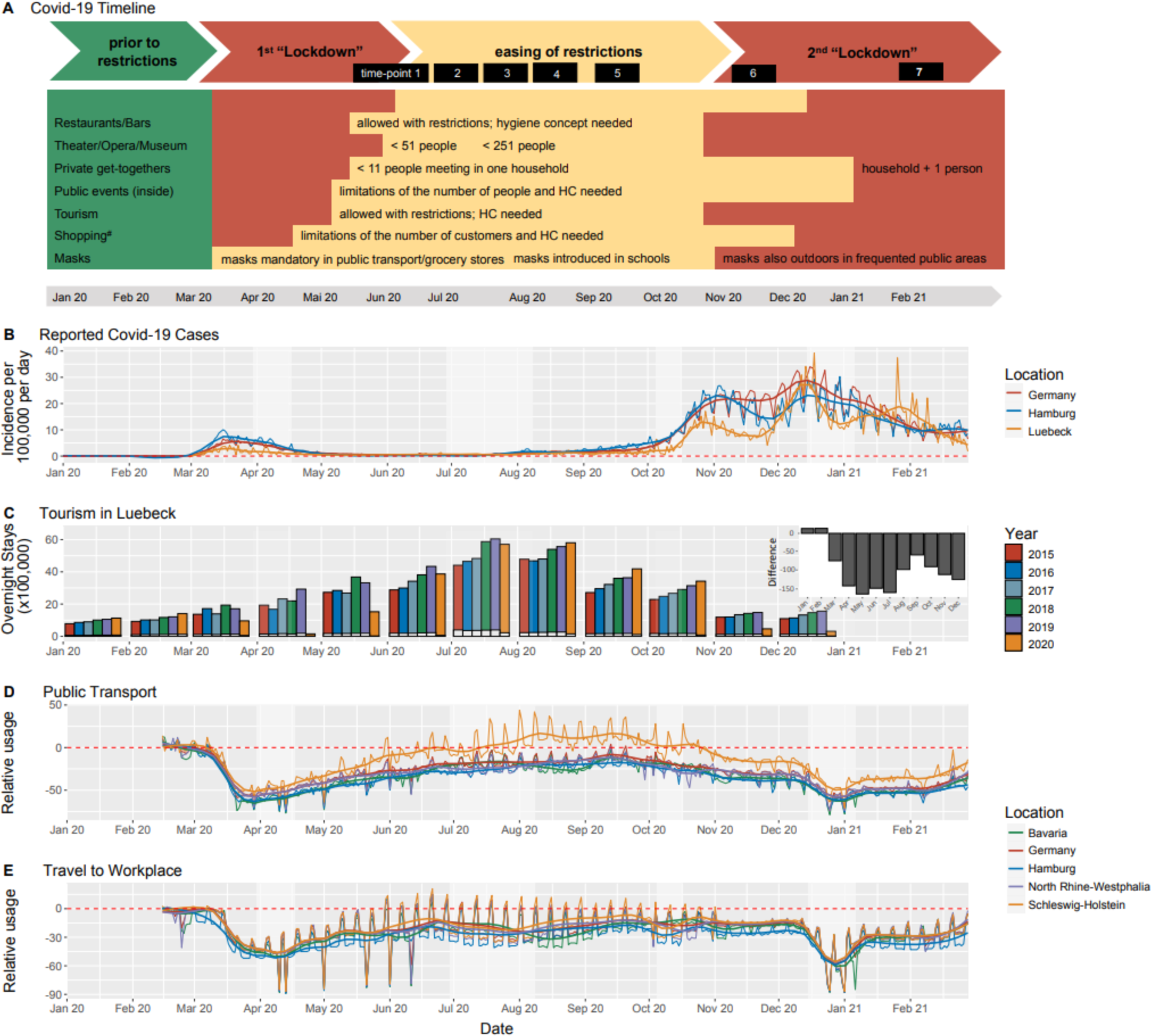
**A**. Lockdown measures in Lübeck, Schleswig-Holstein, Germany; Green: No restrictions prior to the pandemic; Yellow/Orange: restriction/obligations, orange indicates more severe restrictions; Red: forbidden/closed/most severe measures; The different timepoints of the ELISA study are shown in the black boxes; ^#^except grocery stores, bakeries, and gas stations. HC – hygiene concept; Detailed information on lockdown measures as well as references are presented in Table S1. **B**. Reported SARS-CoV-2 infections in Lübeck, Hamburg and Schleswig-Holstein; **C**. Overnight stays in the Lübeck area from the years 2015-2020. Black bars indicate the number of foreign tourists and the difference in overnight stays of foreign tourists in 2020 compared to 2015-2019 (inset). **D. and E**. Usage of public transportation and travel to work in Schleswig-Holstein, Germany and three other German States. Data obtained from Google and reported as relative change from January/February 2020. The thick lines denote a Loess fit to the daily data. The light gray backdrops indicate school holidays in Schleswig-Holstein.

### Study setting

The greater Lübeck area is located on the Baltic Sea coast (**Figure S1**) and a tourist magnet with >2 million overnight stays in 2019 and an average of >50,000 visitors per day with high season in the summer. Until the second wave of the COVID-19 pandemic in Germany, Lübeck was a low-prevalence area (**Figure 2.B**). While there was practically no tourism during the two lockdowns in March and December, tourism in the summer and fall (June to October 2020) reached or even surpassed the already high influx of tourists in previous years (**Figure 2.C**). These numbers were supported by mobility data from Google showing a significant increase in the use of public transportation relative to all of Germany (p-value <10^−30^, moderated F-test) including other touristic hotspots (**Figure 2.D**). Travel to workplaces, however, remained similar across all states and Germany (p-value=0.39; moderated F-test) (**Figure 2.E**).

### Detection of SARS-CoV-2-Infections

Trained study personnel performed deep nasal and oropharyngeal swabs^11^ for RNA isolation and SARS-CoV-2 quantitative Real-Time PCR using validated PCR platforms and accredited SARS-CoV-2 diagnostic pipelines (Supplement;Methods).

For antibody testing, a venous blood sample was drawn. The EUROIMMUN SARS-CoV-2 S1 IgG (#EI 2606-9601-2 G) ELISA was performed according to the manufacturer′s instructions for examination time-points 1, 6, and 7 and partially for time-points 3 and 5 for participants with increased anti-S1 IgG antibody titers at time-point 1. Antibody titers against the S1 antigen of >1.1 were considered positive. Values between 0.8 and 1.1, which are considered borderline, were not included in further analyses.^12^ All persons with antibody-positive results were contacted by telephone to inquire about potential external PCR-positive test results obtained in between study visits.

### App data

Participants were initially asked to answer a baseline questionnaire including demographic data, level of education, symptoms of infection and allergies, profession, travel, activities of daily living, details of adherence to lockdown and later precautionary measures, contact with children, and self-reported results of previous SARS-CoV-2 tests (Supplement;Questionnaires). Participants were asked to repeat this App-based questionnaire (using an adaptation of the MillionFriends App by Perfood GmbH (https://perfood.de/)) every three days until September 2020, corresponding to time-points 1-5. In December 2020 and February 2021, participants were asked to fill out the questionnaire only once (time-points 6 and 7).

### Statistical analysis

Data of the ELISA study was stored in a central study database and was pseudonymized for subsequent analyses. We used frequency tables and cross tabulations to characterize the ELISA cohort and its subgroups. The prevalence of PCR- or antibody-positive participants was estimated using the Kaplan-Meier method. Persons who did not attend all study visits were censored after their last visit. We also censored all vaccinated participants at the last time-point (n=269, 8.8% of the cohort). To explore the potential impact of symptoms on PCR- or antibody-positive results, we marked a person as symptom-positive if a symptom was present for at least one time-point during the study period. For analyzing behavior and quality of life, we calculated a mean behavior score over time for each variable and participant and dichotomized the variables at the median into low/high levels of behavior. We used Cox-Regression to estimate hazard ratios for symptom and behavior variables and calculated 95% confidence intervals for prevalence and risk estimates. Differences in temporal trends were analyzed by a moderated F-test on a regression spline curve with 5 degrees of freedom.

## Results

### Study cohort

The ELISA cohort (n=3051) comprises 56% females and 44% males; the mean age is 45.6 years (SD 15.2 range: 18-79 years). Demographic details of the cohort and its two overlapping subgroups (the high-exposure subgroup includes 523 persons from the population-matched subgroup), and results of the SARS-CoV-2 PCR and antibody testing are summarized in **Table 1**.

**Table 1.** Study participants and group-wise results of antibody screening.

|  |  |  |  |  |
| --- | --- | --- | --- | --- |
|  |  | Value | AB- or PCR positive prevalence |  |
|  |  | N (%) | N | % (CI) |
| **ELISA cohort (total)** |  | **3051 (100)** | 89 | 3.5 (2.8 to 4.2) |
| Gender | Women | 1710 (56) | 45 | 3.1 (2.2 to 4.0) |
|  | Men | 1341 (44) | 44 | 3.9 (2.8 to 5.0) |
| Age | 18-39 | 1199 (39) | 35 | 3.7 (2.5 to 5.0) |
|  | 40-59 | 1164 (38) | 31 | 3.0 (2.0 to 4.1) |
|  | 60-79 | 688 (23) | 23 | 3.6 (2.2 to 5.1) |
| Flagged as high-contact person |  | 1645 (54) | 47 | 3.6 (2.6 to 4.6) |
| Education | low | 916 (30) | 33 | 4.4 (2.9 to 5.9) |
|  | high | 2127 (70) | 55 | 3.0 (2.2 to 3.8) |
| **Population-matched subcohort** |  | **1929 (100)** | 57 | 3.4 (2.6 to 4.3) |
| Gender | Women | 1046 (54) | 17 | 2.3 (1.1 to 3.5) |
|  | Men | 883 (46) | 40 | 4.4 (3.1 to 5.8) |
| Age | 18-39 | 669 (35) | 18 | 3.5 (1.9 to 5.2) |
|  | 40-59 | 656 (34) | 16 | 2.7 (1.4 to 4.0) |
|  | 60-79 | 604 (31) | 23 | 4.1 (2.5 to 5.8) |
| Flagged as high contact person |  | 523 (27) | 15 | 3.8 (1.9 to 5.8) |
| Education | low | 550 (29) | 21 | 4.5 (2.6 to 6.4) |
|  | high | 1375 (71) | 36 | 3.0 (2.0 to 4.0) |
| **High-exposure subcohort** |  | **1645 (100)** | 47 | 3.6 (2.6 to 4.6) |
| Police, army, fire brigade |  | 90 (5) | 4 | 4.5 (0.2 to 8.9) |
| Nurses |  | 236 (14) | 13 | 12.2 (5.0 to 19.0) |
| Other medical staff |  | 41 (2) | 2 | 4.7 (0.0 to 10.9) |
| Sales and distribution |  | 186 (11) | 5 | 3.2 (0.4 to 6.0) |
| Daycare teachers |  | 131 (8) | 5 | 4.2 (0.6 to 7.9) |
| School teachers |  | 160 (10) | 2 | 1.5 (0.0 to 3.6) |
| Students |  | 255 (16) | 9 | 4.2 (1.5 to 7.0) |
| Physician - outpatient practice |  | 122 (7) | 1 | 0.9 (0.0 to 2.7) |
| Physician - hospital |  | 142 (9) | 3 | 2.7 (0.0 to 5.8) |
| Other |  | 282 (17) | 3 | 1.4 (0.0 to 3.0) |

Note: There is overlap of the population-matched and the high-exposure subcohorts with 523 persons from the population-matched group also having high exposure.

### Acute PCR-confirmed SARS-CoV-2 infections and anti-S1 IgG levels

Participation rates were 75% to 98% across all seven time-points (**Figure 1**). Two study participants reported PCR-confirmed SARS-CoV-2 infection before the first examination. The number of individuals with reported PCR-confirmed SARS-CoV-2 infection in the Study App increased over the course of the study and reached n=40 in February 2021. In addition, three and two individuals were tested positive at the study center in November/December 2020 and February 2021, respectively. Twenty-four participants were seropositive at baseline; 61 developed anti-S1 IgG levels above the cut-off over the study course (**Figure 3.A**). In relative numbers, the seropositivity rate within the study cohort increased slightly until September 2020, more pronounced in November/December 2020, and even more in February 2021 (**Figure 3.B**). The frequency of PCR-confirmed SARS-CoV-2 infections showed a similar tendency, however, with a peak in November/December 2020 (**Figure 3.B**).

**Figure 3.**
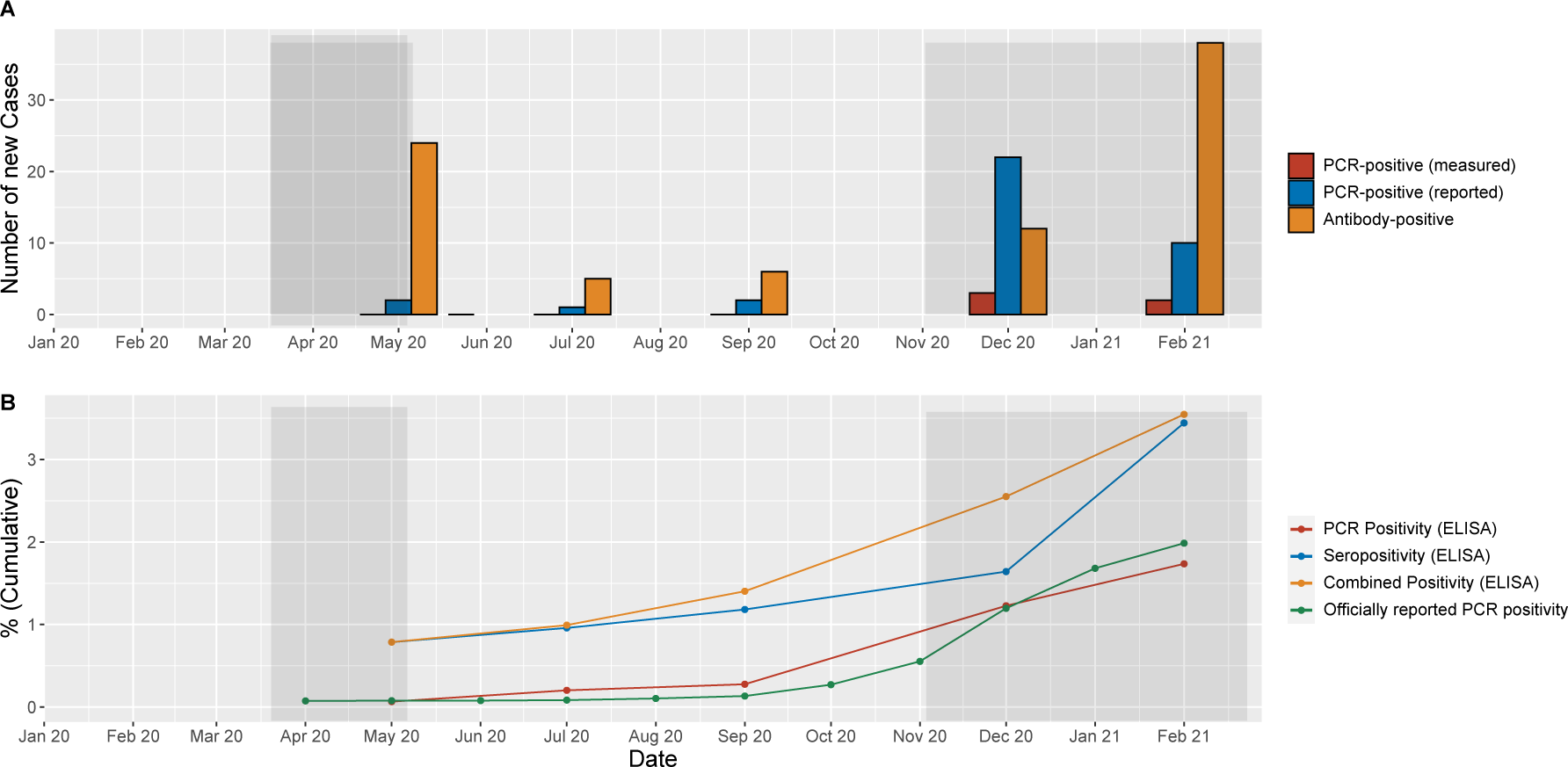
SARS-CoV-2 virus and anti-S1 IgG profiles from May 2020-February 2021, **A**. Number of PCR-confirmed SARS-CoV-2 infections and individuals with anti-S1 IgG > 1.1 per timepoint. PCR-confirmed infections are shown in orange (reported in the Study App) and red (positive PCR test at the study center), respectively. Newly detected anti-S1 IgG positive cases are depicted in blue. **B**. Frequency of PCR-confirmed infections within the study compared to officially reported results and anti-S1 IgG positivity rates. Data are presented in a cumulative fashion, whereby study participants were classified as positive at all future timepoints starting with their first positive test. Officially reported frequency of positive PCR results is shown in black (https://npgeo-corona-npgeo-de.hub.arcgis.com, rates of the last day of the month in Lübeck are shown). PCR positivity (reported and tested) at the study center is depicted in red, the total frequency of positive antibody results within the study cohort in blue, and combined positivity (PCR-confirmed infections and positive anti-S1 antibodies) in green. The gray bars on the right- and the left-hand side of each panel indicate lockdown periods.

The stability of anti-S1 IgG antibody titers over time is depicted in **Figure S3**. Within the one-year observation period, 89 persons (3.5%) were antibody-positive (n=85) and/or had a positive PCR test result (n=42) (**Table S2**). Thirty-nine of the 42 PCR-positive persons had an additional antibody test after having tested positive before; 37 (95%) of them were positive for Anti S1 IgG antibodies. The five and six anti-S1 IgG antibody-positive samples at time-point 3 and 5, respectively, had shown increased antibody titers at baseline, however, with ratios <1.1. Prevalence of PCR- or seropositivity in the population matched subcohort was almost 2-fold higher in men than in women (4.4% vs. 2.3%) and increased risk was detected in several high-exposure groups. The highest prevalence of 12.2% was observed for nurses, followed by other medical staff, police, army, firemen, and students. Assuming that antibody-positive findings represent the true prevalence, about 92% of all infections were missed by PCR testing in May 2020 (n=22/24). By February 2021, only 29% of the new antibody positive cases (11/38) remained undiagnosed.

### Analysis of risk stratification, behavior, quality of life and correlation with antibody profiles

The development of symptoms, behavior, and quality of life during the pandemic (**Figure 4.A**) were assessed from April 2020 to February 2021 and are based on self-reports from 3051 study participants in 90,923 electronic questionnaires.

**Figure 4.**
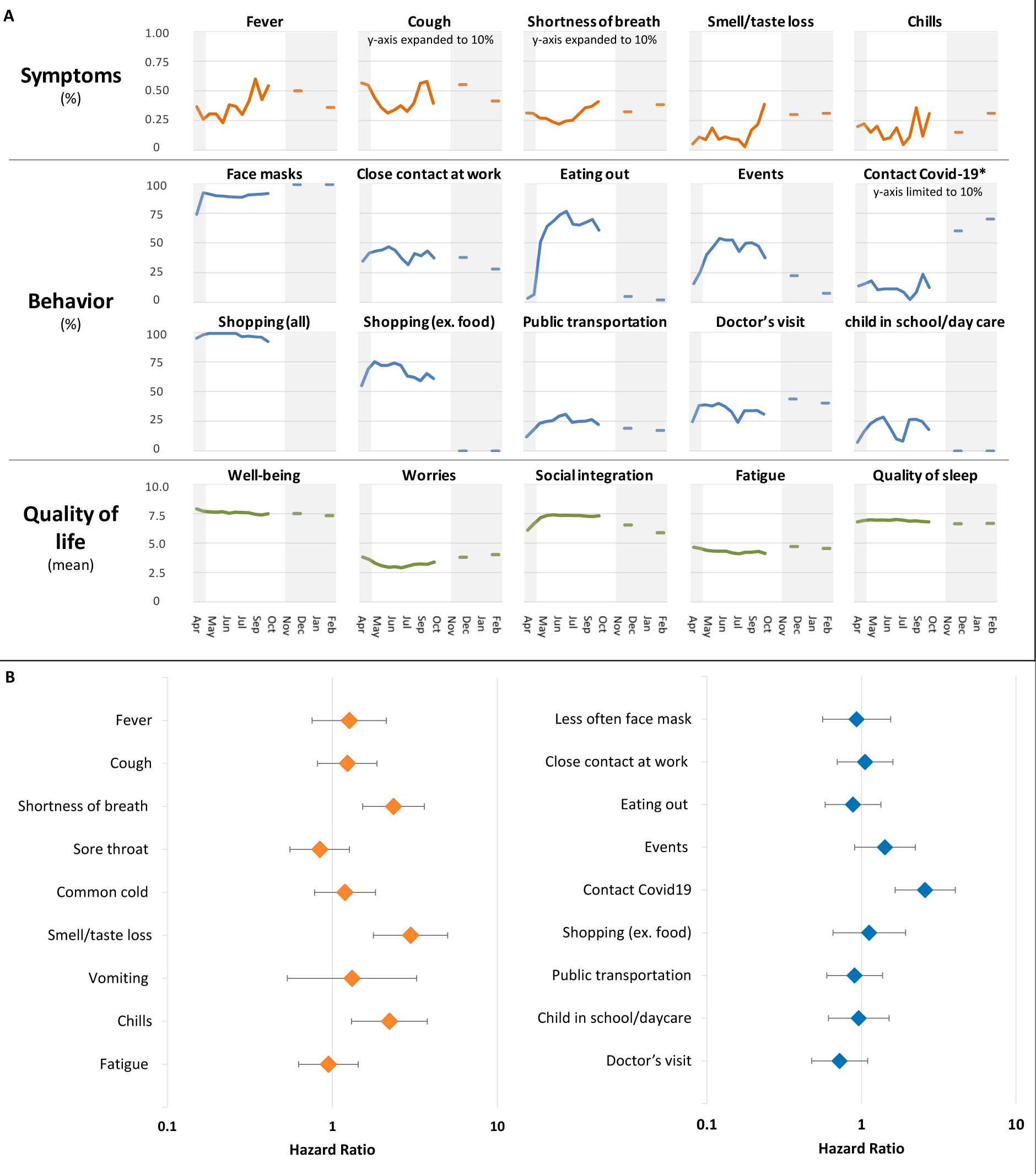
**A**. Symptoms (orange), behavior (blue), and quality of life (green) during the pandemic from April 2020 to February 2021, aggregated by two-week intervals. Symptoms and behavior are shown as percent values. The y-axis is scaled to 1% for symptoms, 100% for behavior, and for some variables set to 10% as indicated; quality-of-life variables are shown as mean value of a score ranging from 1-10 (1 lowest, 10 highest). The gray bars on the right- and the left-hand side of each panel indicate lockdown periods. At examination time-points 6 and 7, questionnaires were no longer obtained every three days but only at two individual time-points each. **B**. Forest plot depicting hazard ratios for antibody- or PCR-positive results for selected symptoms (left) and behaviors (right).

Frequencies of infection symptoms rose between lockdown periods and dropped during the second lockdown. Behaviors were either stable (e.g., use of face masks), increased in frequency between lockdowns in relation to alleviation of the lockdown measures (e.g., eating out) and dropped again sharply during the second lockdown. The dip in frequency of many behaviors in the summer is related to the 6-week summer vacation period. Contact to COVID-19 patients increased during the second lockdown. Quality of life remained stable for all included parameters with the exception of a moderate drop in social integration during the second lockdown period (**Figure 4.A**).

The occurrence of three of the self-reported symptoms was associated with anti-S1 IgG antibody/PCR positivity: loss of smell/taste (HR 2.98 [95%CI 1.78 to 4.99]), shortness of breath (HR 2.35 [95%CI 1.53 to 3.61]), and chills (HR 2.21 [95%CI 1.30 to 3.76]) (**Figure 4.B, Table S3**). Of note, predictive value of these symptoms was low (e.g., only 18 out of 244 (7.4%) persons with impairment of smell or taste were infected). Analysis of different behaviors revealed “contact to persons with COVID-19” as the most relevant factor for testing positive (HR 2.58 [95%CI 1.65 to 4.04]) (**Figure 4.B**). Other factors, such as frequent use of public transportation, shopping or close contacts at work did not impact infection rates. Mobility data revealed an association between the decrease of the use of public recreation areas-likely indicating reduced outdoor activities after the summer - and the increase of SARS-CoV-2 infections in the study area (**Figure S3**).

## Discussion

The ELISA Study is the first to provide longitudinal data throughout the SARS-CoV-2 pandemic on a prospectively followed, population-based cohort. Compared to the earlier point-of-care test approaches,^13^ it is further unique for its comprehensive design of seven rounds of PCR and antibody testing, combined with close digital tool-based recording of symptoms, behavior, and well-being. There is a critical need to identify specific groups and situations associated with an increased risk for SARS-CoV-2 infection,^14^ as such knowledge is urgently needed to inform choice and duration of preventive measures. Currently, only modeling and cross-sectional studies with relatively small sample sizes are available.^15^ Moreover, in most countries, including Germany, no structured population-based measures of high methodological quality have been implemented as yet, and data assessed by the health authorities still remain to be of insufficient quality. The high retention rate and acceptance among our study participants over a one-year period confirm feasibility of large-scale, long-term surveillance studies. As monitoring of 1% of the local population successfully mirrored reported infection rates of the region with acceptable precision, our study design may serve as an efficient and cost-effective model.

Despite a large influx of tourism and mobility in the summer, as well as an unexpected rise of infection rates in the previously low-incidence study region in the fall/winter 2020/2021, overall antibody seropositivity remained low across all study time-points and profoundly lower compared to high-prevalence areas such as Tirschenreuth (Germany),^16^ Val Gardena (Italy),^17^ or New York City (USA).^18^ Seropositivity in the Lübeck study area reached only about one twentieth of the ∼70% required for herd immunity further stressing the vulnerability of the vast majority of the population and the pivotal role of vaccination. Of further note is the dramatic decrease of the proportion of undetected SARS-CoV-2 infections from 92% in Mai 2020 to 29% in February 2021 which is due to vastly increased testing efforts. At the beginning of the pandemic, the true incidence may have been up to 11 times higher than the official figures, confirming early estimates.^19^

Several COVID-19-linked symptoms,^20^ infection risk-associated behaviors, and higher occupational exposure in health care personnel,^21^ police, fire-brigade, and students were confirmed or newly identified as plausible risk factors for SARS-CoV-2 infection. In contrast, other potential reported risk factors did not seem to contribute to the risk of infection, such as use of public transportation,^7^ eating out^22^ or having a child in school.^10,23^ Another important area of discussion relates to schools as a relevant driver of the pandemic. In our study, only 2/160 teachers were antibody-positive after one year, confirming results from a prospective study conducted in primary schools in England.^24^ Other studies have used mobility networks to inform reopening^25^ and identified selected ethnicities^26^ and individuals with close household contacts^27,28^ to be at higher risk of infection.

Despite the massive influx of tourism from Germany into the Lübeck area in the summer of 2020 in parallel with an almost complete ease of lockdown restrictions and high mobility of the study population itself, we did not observe an increase in infection rates suggesting that easing of measures was safe. Notably, all of these findings have to be interpreted in the context of (partial) lockdown periods, continuous hygiene measures, and the low prevalence in our study area. However, despite limited interpretability of our results by the small size of individual subgroups and low predictive values, we do provide important data to inform the ongoing discussion of lockdown measures, as well as of prioritization of vaccination of specific at-risk groups.

The sharp rise in infection rates in late 2020 raises the important question as to whether we could have predicted the ‘second wave’. Not surprisingly, contact with infected individuals was the strongest sole risk factor for SARS-CoV-2 infection in our study. We hypothesize a fast rise in diffuse transmission, as soon as incidences surpass a critical threshold and quickly reach exponential growth when no (new) lockdown measures are promptly imposed. While there were no changes of behavior or emerging differences in our high-exposure group heralding an imminent ‘second wave’, we noted a strong anti-correlation between rise in the number of infections in Lübeck and the use of public recreation areas, likely indicative of more indoor activities that may have fostered contact with infected individuals. Strengths of our study include its comprehensive and longitudinal design, the quality of virus and antibody testing, exceptionally high cohort retention, and the dynamic nature of the study region in terms of tourism, mobility and sudden change from a low-to a high-prevalence region. Limitations are overrepresentation of highly motivated study participants with an above-average education level and the risk of false-positive test results in a low-incidence setting. The latter is mitigated by the conservatively chosen cut-offs for antibody positivity, consistent results over several time-points, and high concordance between PCR and antibody positivity.

In conclusion, we i) provide a model for effective, regional surveillance of the pandemic; ii) underscore the role of vaccination especially in low-prevalence regions remaining far from herd immunity; iii) identify plausible infection risk factors informing public health measures; iv) demonstrate that easing of lockdown measures appears safe even over several months at times of low prevalence rates; v) underline that continuous monitoring of infections and immediate and consequent imposition of appropriate measures are mandatory to contain virus spread in case of an incipient rise in infection rates.

## Data Availability

All data are available from the corresponding authors upon request.

## Acknowledgments

We are very grateful to the participants for taking part in our study over the entire one-year period, strengthening our results through the high cohort retention rate. Moreover, we would like to thank Aiham Alabid, Sylwia Dankert, Katja Herrmann-Malzahn, Isabell Rosin, and Christoph Westenberger for excellent assistance in creating the website and organizing the study, and Karolin Ratalics, Nina Janzen, and Torsten Schröder for technical support with the study app. Finally, we thank Vera von Kopylow, Inga Künsting, Anne S. Lixenfeld, and Emily L. Martin for performing antibody analyses.

## Funding

The study was funded by the Federal Ministry of Education and Research (BMBF) within the Sub-project B-FAST of the joint project National Research Network of University Medicine on COVID-19 (BMBF - FKZ 01KX2021), by the State of Schleswig-Holstein, by the Tuberculosis Foundation SH, and crowdfunding from the citizens of Lübeck organized by the University of Lübeck.

## Notes

### Competing Interest Statement

The authors have declared no competing interest.

### Funding Statement

The study was sponsored by the Federal Ministry for Education and Research (BMBF), the State of Schleswig-Holstein, by the Tuberculosis Foundation SH, by the University of Luebeck, and by a crowd-funding campaign organized by the University of Luebeck.

### Author Declarations

IRB of the University of Luebeck The ELISA (Luebeck Longitudinal Investigation of SARS-CoV-2 Infection) study was approved by the ethics committee of the University of Luebeck (Az. 20-150)

