## Supplement for "Cohort-based surveillance of SARS-CoV2 transmission mirrors infection rates at the population level: a one-year longitudinal study"

### Supplementary Appendix

#### Table of Contents


Supplement – Methods page 1

Supplement – Tables page 3

Supplement – Figures page 6

Supplement – Questionnaires page 8

Supplement – References page 21

### Supplement – Methods

#### Real-Time PCR SARS-CoV-2 analysis

Medical students performed deep nasal and oropharyngeal swabs under supervision of medical doctors and combined them into a single tube for each participant prior to RNA isolation. SARS-CoV-2 quantitative Real-Time PCR (RT-PCR) from dry swabs was performed at the National Reference Center for Mycobacteria at the Research Center Borstel (RCB), Leibniz Lung Center[1] and the University Hospital Schleswig-Holstein (UKSH), Lübeck, using validated PCR platforms. External supporting sites included the LADR Central Lab Dr. Kramer & Colleagues, Geesthacht and Centogene GmbH, Rostock,[2] using accredited SARS-CoV-2 diagnostic pipelines. Briefly, dry swabs were suspended within 24 hours and processed for nucleic acid extraction using QIAamp® Viral RNA Kits (Qiagen, Hilden) or the NucliSens® easyMAG™ platform (Biomérieux), according to the manufacturers' instructions. Nucleic acids were eluted and quantitative RT-PCR was performed according to the manufacturers' instructions using the first WHO Emergency Use[3] listed In vitro diagnostics coronavirus (COVID-19) genesig® Real-Time PCR assay (Primerdesign™ Ltd), or the iTaq Universal Probes 1-Step kit (Bio-Rad, Feldkirchen) in conjunction with primers and probes from the SARS-CoV-2 (2019-nCoV) CDC qPCR Probe Assay (IDT, Leuven, Belgium). All in-house PCR assays were validated and passed the interlaboratory quality control (INSTAND, Düsseldorf).

#### Supplement – Tables

**Table S1.** Lockdown measures and ease of restrictions in Schleswig-Holstein, Germany, March 2020 to February 2021

| Date | Most important measure | Schools/Daycare | Private meetings | Events | Shopping# | Bars/ Restaurants | Theater/ Opera/ Museum | Tourism | Reference |
|---|---|---|---|---|---|---|---|---|---|
| 2020/03/17 | First-time implementation of restrictions of daily life | closed | not allowed | not allowed | closed | closed | closed | not allowed | Decree by the federal state government of Schleswig-Holstein, "SARS-CoV-2-Bekämpfungsverordnung (SAR-CoV-2-BekämpfVO)", March 17, 2020 |
| 2020/04/08 | Detailed provided about private gatherings by the authorities | closed | <11 people | not allowed | closed | closed | closed | not allowed | Decree by the federal state government of Schleswig-Holstein, "SARS-CoV-2-Bekämpfungsverordnung (SAR-CoV-2-BekämpfVO)", April 8, 2020 |
| 2020/04/24 | Face masks introduced in public transport and grocery stores | closed | <11 people | not allowed | closed | closed | closed | not allowed | Decree by the federal state government of Schleswig-Holstein, "SARS-CoV-2-Bekämpfungsverordnung (SAR-CoV-2-BekämpfVO)", April 24, 2020 |
| 2020/05/01 | Reopening of shops with a size below 800 $m^2$ | closed | <11 people | <51 people | <800$m^2$, R, HC | closed | closed | not allowed | Decree by the federal state government of Schleswig-Holstein, "SARS-CoV-2-Bekämpfungsverordnung (SAR-CoV-2-BekämpfVO)", May 1, 2020 |
| 2020/05/16 | Reopening of restaurants and hotels with restrictions | closed | <11 people | <51 people | R, HC | R, HC | closed | R, HC | Decree by the federal state government of Schleswig-Holstein, "SARS-CoV-2-Bekämpfungsverordnung (SAR-CoV-2-BekämpfVO)", May 16, 2020 |
| 2020/06/05 | Reopening of theaters, opera, museums with restrictions | closed | <11 people | <251 people | R, HC | R, HC | <101 people, HC | R, HC | Decree by the federal state government of Schleswig-Holstein, "SARS-CoV-2-Bekämpfungsverordnung (SAR-CoV-2-BekämpfVO)", une 5, 2020 |
| 2020/06/08 | Reopening of primary schools | primary schools open | <11 people, | <251 people | R, HC | R, HC | <101 people, HC | R, HC | https://www.tagesschau.de/inland/coronakrise-lockerungen-laender-101.html |
| 2020/07/15 | Further ease of restrictions | primary schools open | <11 people | <501 people | R, HC | R, HC | <251 people, HC | R, HC | Decree by the federal state government of Schleswig-Holstein, "SARS-CoV-2-Bekämpfungsverordnung (SAR-CoV-2-BekämpfVO)", July 15, 2020 |
| 2020/08/10 | Reopening of all schools after summer vacation | open | <11 people | <501 people | R, HC | R, HC | <251 people, HC | R, HC | https://schleswig-holstein.de/DE/Schwerpunkte/Coronavirus/Schulen_Hochschulen/corona_schule.html#docb5a355f7-c9fa-4b6d-86f9-452954b9875abodyText6 |
| 2020/08/24 | Masks also required at schools | open | <11 people | <501 people | R, HC | R, HC | <251 people, HC | R, HC | Decree by the federal state government of Schleswig-Holstein, "SARS-CoV-2-Bekämpfungsverordnung (SARS-CoV-2-BekämpfV)", August 22, 2020 |

| | | | | | | | | | |
|---|---|---|---|---|---|---|---|---|---|
| 2020/11/01 | Aggravation of restrictions due to severely increasing cases, masks required also outdoors in highly frequented public places | partly open | <11 people | <101 people | R, HC | closed | closed | not allowed | Decree by the federal state government of Schleswig-Holstein, “SARS-CoV-2-Bekämpfungsverordnung (SARS-CoV-2-BekämpfV)”, November 1, 2020 |
| 2020/11/29 | Schools nearly completely closed between December 16, 2020 and January 9, 2021 | almost completely closed | <11 people | <101 people | R, HC | closed | closed | not allowed | Decree by the federal state government of Schleswig-Holstein, “SARS-CoV-2-Bekämpfungsverordnung (SARS-CoV-2-BekämpfV)”, November 29, 2021 |
| 2020/12/14 | Closure of all stores except for food and hygiene supplies, no events allowed, special restrictions for Christmas and New Year's Eve 2021 holidays. | almost completely closed | <5 people | not allowed | closed | closed | closed | not allowed | Decree by the federal state government of Schleswig-Holstein, “SARS-CoV-2-Bekämpfungsverordnung (SARS-CoV-2-BekämpfV)”, December 12, 2020 |
| 2021/01/08 | Further limitation of private meetings (only one additional person to each household), extension of school closings | almost completely closed | household + one other person | not allowed | closed | closed | closed | not allowed | Decree by the federal state government of Schleswig-Holstein, “SARS-CoV-2-Bekämpfungsverordnung (SARS-CoV-2-BekämpfV)”, January 8, 2021 |

#except grocery stores, gas stations, bakeries. R – Restrictions; HC – Hygiene concept needed; All decrees of the federal state government of Schleswig-Holstein are stored on the following website: https://www.schleswig-holstein.de/DE/Landesregierung/IV/Service/GVOBl/gvobl_node.html; All websites were accessed and the decrees were downloaded on May 4, 2021.

**Table S2.** **PCR and antibody (AB) positive cases**

| | PCR⁺ or AB⁺ | | PCR⁺ | | AB⁺ | | | | |
|---|---|---|---|---|---|---|---|---|---|
| | at risk n | events n (%) | at risk n | events n (%) | at risk n | events n (%) | PCR⁺ before AB⁺ n (%) | missed by PCR n (%) | Ratio detected missed |
| May 2020 | 3051 | 24 (0.79%) | 3051 | 2 (0.07%) | 3051 | 24 (0.79%) | 2 (8.3%) | 22 (91.7%) | 1:11 |
| Jul 2020 | 2894 | 5 (0.17%) | 2923 | 1 (0.03%) | 2894 | 5 (0.17%) | 1 (20.0%) | 4 (80.0%) | 1:4 |
| Sep 2020 | 2659 | 8 (0.30%) | 2701 | 2 (0.07%) | 2665 | 6 (0.23%) | 0 (0.0%) | 6 (100.0%) | - |
| Dec 2020 | 2580 | 29 (1.12%) | 2613 | 25 (0.96%) | 2581 | 12 (0.46%) | 7 (58.3%) | 5 (41.7%) | 1:0.7 |
| Feb 2021 | 2057 | 23 (1.12%) | 2098 | 12 (0.57%) | 2073 | 38 (1.83%) | 27 (71.1%) | 11 (28.9%) | 1:0.4 |
| Total | | 89 (3.46%) | | 42 (1.69%) | | 85 (3.44%) | 37 (43.5%) | 48 (56.5%) | 1:1.3 |

For column 'AB⁺ or PCR⁺' an event was assigned to the time of the first occurrence of positive PCR or antibody test. Column 'PCR⁺': 39 of 42 PCR⁺ had AB measured after positive PCR Test. 37 of them were AB⁺ and 2 were AB⁻ at the end of the study. Of the 3 remaining PCR⁺ cases, 1 case was lost to follow up, 2 had been vaccinated before AB measurement.

**Table S3.** **Hazard ratios (HR) for a PCR or antibody (AB) positive result for selected symptoms and behaviors**

| | | PCR or AB positive | |
|---|---|---|---|
| | N (%) | N (%) | HR (95%CI) |
| **Symptoms** | | | |
| Fever | 529 (17.3) | 18 (3.4) | 1.27 (0.75 to 2.12) |
| Cough | 1389 (45.5) | 45 (3.2) | 1.23 (0.81 to 1.87) |
| Shortness of breath | 618 (20.3) | 33 (5.3) | 2.35 (1.53 to 3.61) |
| Sore throat | 1657 (54.3) | 44 (2.7) | 0.84 (0.55 to 1.27) |
| Common cold | 1716 (56.2) | 54 (3.1) | 1.19 (0.78 to 1.82) |
| Smell/taste loss | 244 (8.0) | 18 (7.4) | 2.98 (1.78 to 4.99) |
| Vomiting | 137 (4.5) | 5 (3.6) | 1.31 (0.53 to 3.24) |
| Chills | 312 (10.2) | 17 (5.4) | 2.21 (1.30 to 3.76) |
| Fatigue | 1523 (49.9) | 43 (2.8) | 0.95 (0.62 to 1.43) |
| **Behaviors** | | | |
| Less face masks | 698 (22.9) | 19 (2.7) | 0.93 (0.56 to 1.55) |
| Close contact at work | 1504 (49.3) | 44 (2.9) | 1.06 (0.70 to 1.60) |
| Eating out | 1497 (49.1) | 42 (2.8) | 0.88 (0.58 to 1.34) |
| Events | 691 (22.6) | 27 (3.9) | 1.42 (0.90 to 2.23) |
| Contact COVID-19 | 476 (15.6) | 28 (5.9) | 2.58 (1.65 to 4.04) |
| Shopping (ex. food) | 533 (17.5) | 16 (3.0) | 1.12 (0.65 to 1.93) |
| Public transportation | 1524 (50.0) | 42 (2.8) | 0.90 (0.60 to 1.37) |
| Child in school/daycare | 956 (31.3) | 27 (2.8) | 0.96 (0.61 to 1.51) |
| Doctor's visit | 1532 (50.2) | 39 (3.2) | 0.72 (0.48 to 1.10) |

#### Supplement – Figures

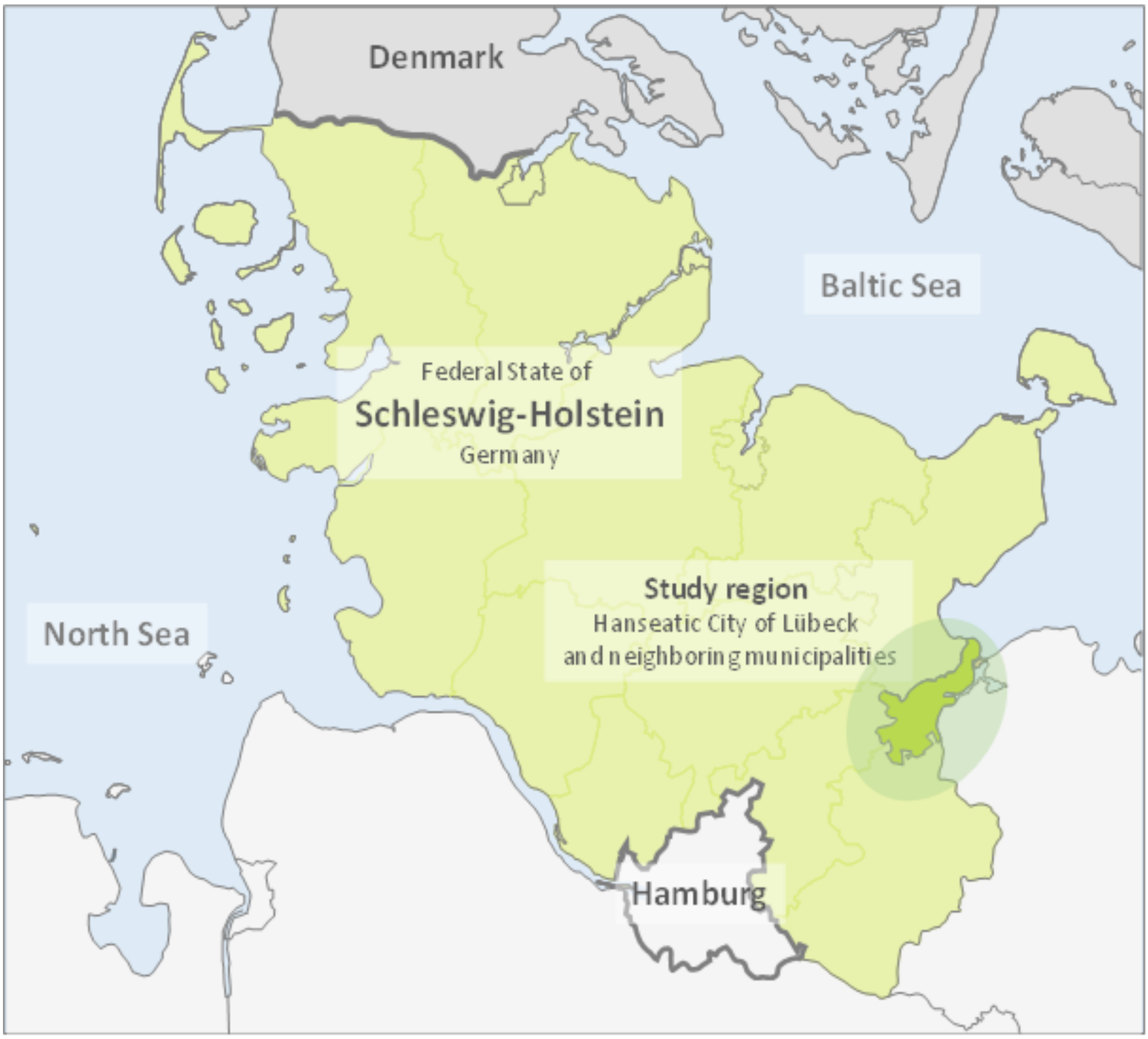


**Figure S1. Study region "greater Lübeck area" (darker green), located in Schleswig-Holstein, Germany.** Lübeck's geographic position is in the north of Germany between the high-prevalence area of the Hamburg and the Baltic Sea coast, a major tourist area.

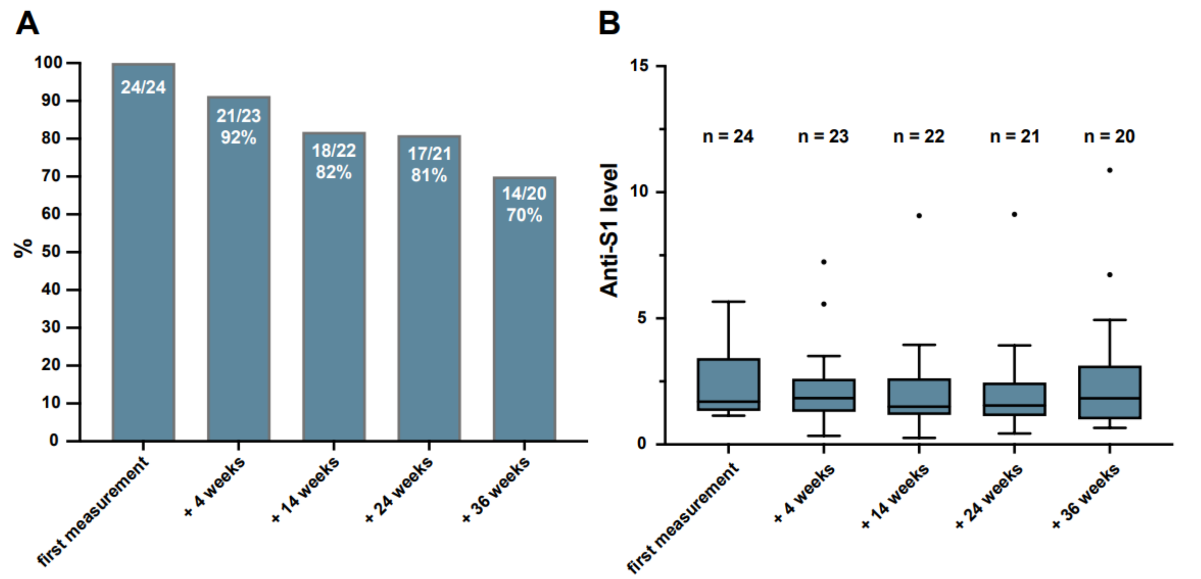


**Figure S2.** **Antibody titer stability over the study course.** A. Out of 24 individuals with anti-S1 IgG >1.1 at baseline, the majority was available for follow-up examination. Individual antibody levels decreased below the cut-off in a proportion of individuals. The bars depict the percentage of participants still reaching anti-S1 IgG > 1.1 four, 14, 24, and 36 weeks after baseline measurement, respectively. B. Median of anti-S1 IgG levels of individuals considered antibody-positive at baseline are shown over time. Data are presented as box and whisker plots and the box extends from the 25th to the 75th percentile. The line in the middle of the box represents the median and Tukey whiskers are used.

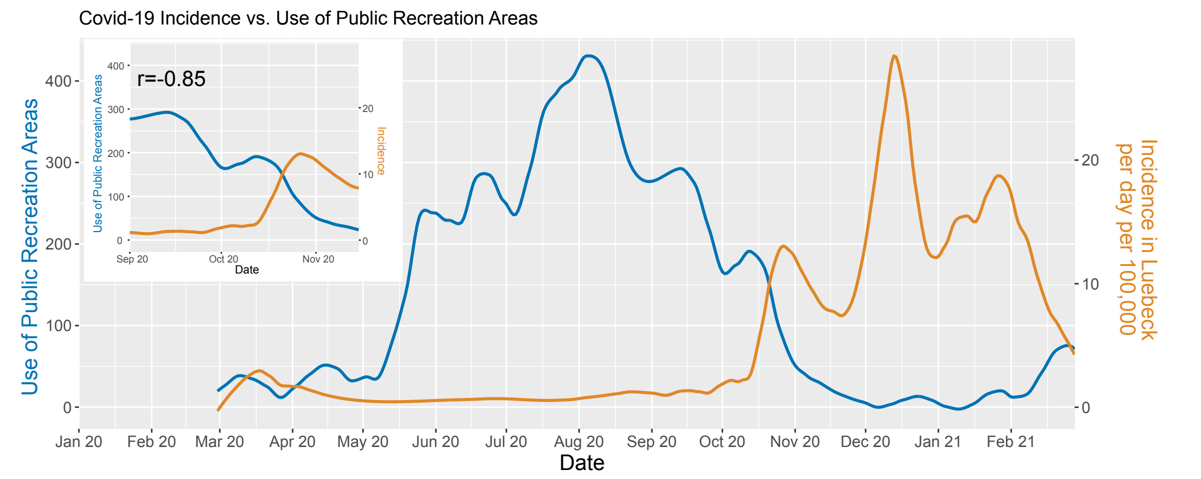


**Figure S3.** **Reverse correlation between the use of public recreation areas (mobility data) and SARS-CoV-2 infections in the study area.** The blue and orange lines depict the Loess-smoothed "Use of Public Recreation Areas" in Schleswig-Holstein based Google Mobility Data and the number of reported Covid-19 infections in Lübeck over the time-course of March 2020 till March 2021. The top-left inset depicts the time from Sep. 1$^{st}$ – Nov. 15$^{th}$ during which the use of public parks declined greatly, and incidences rose in an highly-anti-correlated manner (r=-0.85, Pearson Correlation). The mobility data is based on the relative change relative to the 5-week period Jan 3 – Feb 6, 2020 and has been obtained from *google.com/covid19/mobility/,* accessed on March 31$^{st}$ 2021. Loess smoothing is based on 10% of data points.

#### Supplement – Questionnaires

##### Initial screening questionnaire (prior to study enrolment)

Sex
0. male
1. female

Current weight

Date of birth

Height

Please enter the postal code of the place where you currently live

Which of the following symptoms do you currently have?
0. none
1. fever / increased temperature >37.5 °C
2. cough
3. shortness of breath
4. muscle / limb pain
5. sore throat
6. cold
7. new onset of loss or change of smell
8. new onset of loss or change of taste
9. headache
10. nausea
11. vomiting
12. diarrhea
13. increased sweating
14. chills
15. seizures
16. fatigue/increased tiredness
17. dizziness
18. unsteadiness of stance and/or gait
19. other (free text)

Have you taken your temperature?

Please specify the measured body temperature

Where did you take your temperature?
0. in the mouth
1. in the armpit

2. in the anus
3. in the ear
4. in the groin

Do you have any sputum?

What does the sputum look like?
0. colorless/clear
1. whitish/mucous
2. yellowish/greenish
3. sanguineous
4. other (free text)

When do you experience shortness of breath?
0. during exertion
1. at rest
2. while lying down

Is your nose blocked?

Do you have a runny nose?

What does the nasal discharge look like?
0. colorless/clear
1. whitish/mucous
2. yellowish/greenish
3. sanguineous
4. other (free text)

In the last 72 hours, how often did you vomit?

In the last 72 hours, how often did you have bowel movements?

You have indicated that you currently have other symptoms. Please specify

When did your current symptoms start?

Apart from possible current signs of illness: Which of the following symptoms were present between February 2020 and now?
0. none
1. fever / increased temperature >37.5 °C
2. cough
3. shortness of breath
4. muscle / limb pain
5. sore throat

6. cold
7. new onset of loss or change of smell
8. new onset of loss or change of taste
9. headache
10. nausea
11. vomiting
12. diarrhea
13. increased sweating
14. chills
15. seizures
16. fatigue/increased tiredness
17. dizziness
18. unsteadiness of stance and/or gait
19. other (free text)

When did the symptoms first appear?
0. first half of February
1. second half of February
2. first half of March
3. second half of March
4. first half of April
5. second half of April
6. first half of May
7. second half of May

Have you been in contact with one or more COVID-19 patients who tested positive?

When did the contact occur?
0. first half of February
1. second half of February
2. first half of March
3. second half of March
4. first half of April
5. second half of April
6. first half of May
7. second half of May

Have you ever been tested for Corona virus infection?

Have you been tested once or more than once for Corona virus infection?
0. once
1. several times

When was the testing done?

When did you take your first test for Corona virus infection?

When did you take your last test for Corona virus infection?

Was the test/any of the tests positive for Corona virus?

Have you been hospitalized since January 2020?

When were you admitted to the hospital?

Are you currently still hospitalized?

When were you discharged from the hospital?

Is or was the hospitalization due to a due to COVID-19 disease?

Did you require oxygen during your stay?

Did you need ventilation?

Have you been abroad between February 2020 and today?

Which country did you travel to?
0. Austria
1. Switzerland
2. France
3. Spain
4. Italy
5. USA
6. China
7. South Korea
8. other (free text)

When did you last travel?
0. first half of February
1. second half of February
2. first half of March
3. second half of March
4. first half of April
5. second half of April
6. first half of May
7. second half of May

The next question is about travel within Germany: Have you been outside Schleswig-Holstein between February 2020 and today?

Have you been in one or more of the following Federal States?
0. Hamburg
1. Berlin
2. Bavaria
3. Baden-Wuerttemberg
4. North Rhine-Westphalia
5. other (free text)

When were you last outside Schleswig-Holstein?
0. first half of February
1. second half of February
2. first half of March
3. second half of March
4. first half of April
5. second half of April
6. first half of May
7. second half of May

Specify your highest school degree
0. elementary school
1. secondary school
2. secondary school
3. high school diploma
4. no school degree

unHave you graduated from a university?

Do you belong to one of the following groups or
do you work in one of the following professions?
0. no, I do not belong to any of the following groups
1. geriatric nurse (in nursing home)
2. geriatric nurse (outpatient care service)
3. member of the federal armed forces/federal police force
4. doctor in hospital
5. pharmacist
6. postman
7. bus driver
8. day care teacher
9. hairdresser
10. family doctor
11. hotel employee
12. school teacher
13. employee in delivery service
14. medical specialist in private practice

15. nurse in hospital
16. policeman
17. pensioner or retiree
18. university student
19. waiter
20. other hospital staff (e.g. physiotherapist, dietician, cleaner)
21. school students
22. supermarket staff
23. gas station employee
24. drugstore salesperson

Please enter your profession

In which occupation did you work before you retired?

How many persons live in your household?

How many persons over 60 years of age live in your household?

How many people under the age of 18 live in your household?

How many people in your household are employed?

Do you suffer from allergies?

Which allergies do you suffer from exactly?
0. chemicals
1. house dust/mite
2. hay fever
3. insect venom
4. cow milk
5. crustaceans/seafood
6. latex
7. lupine
8. medicines
9. metals
10. nuts/peanuts
11. fruits/vegetables/grains
12. perfume/cosmetics
13. sulfur dioxide and sulfites
14. mustard
15. sesame
16. animal hair
17. I have allergies, but I don't know which ones
18. other (free text)

Do you currently suffer from allergy symptoms (watery eyes, runny nose?

Do you have one or more of the following pre-existing conditions?
0. none
1. asthma
2. high blood pressure
3. COPD (chronic obstructive pulmonary disease)
4. depression
5. diabetes
6. inflammatory bowel disease (Crohn's disease, ulcerative colitis)
7. inflammatory diseases of the skin (e.g. psoriasis)
8. elevated blood lipid levels
9. fatty liver
10. heart attack in the past
11. frequent urinary tract infections
12. heart failure (cardiac insufficiency)
13. osteoporosis
14. cancer
15. pneumonia in the past
16. migraine
17. Parkinson's disease
18. multiple sclerosis
19. frequent sinusitis (inflammation of the sinuses)
20. renal insufficiency (impaired kidney function)
21. irritable bowel syndrome
22. rheumatic disease of the joints
23. rheumatic disease of internal organs/vessels
24. hypothyroidism
25. stroke in the past
26. celiac disease
27. other (free text)

What type of pathogen caused your pneumonia?
caused your pneumonia?
0. viruses (e.g. influenza viruses)
1. bacteria (e.g. pneumococci)
2. fungi
3. do not know

What medications do you take regularly?
0. none
1. antidiabetics (e.g. metformin, insulin)
2. antibodies/biologics (e.g. Humira, MabThera, Remicade)
3. asthma inhalers (e.g., sultanol, budenoside)

4. antihypertensives (e.g. metoprolol, enalapril)
5. cortisone (e.g. prednisolone, hydrocortisone)
6. inhibition of blood clotting (e.g. ASS, Clopidogrel)
7. "strong" blood thinners (e.g. Marcumar, Xarelto, Eliquis)
8. immunosuppressants (e.g., azathioprine, methotrexate)
9. lipid-lowering drugs (e.g. simvastatin, pravastatin)
10. thyroid tablets (e.g. L-thyroxine, thyrostatic drugs)
11. painkillers: opiates (morphine, tramadol)
12. painkillers: non-steroidal anti-inflammatory drugs (e.g. ASS, Ibuprofen, Diclofenac)
13. other (free text)

Do you take any food additives?
0. none
1. vitamin D3
2. vitamin C
3. vitamin B1
4. vitamin B3
5. vitamin B12
6. vitamin K
7. vitamin E
8. multivitamin
9. magnesium
10. zinc
11. selenium
12. silica
13. other (free text)

What is the name of the multivitamin supplement you are taking?

Have you ever received one or more of the following of the following vaccinations?
0. none
1. tuberculosis (BCG-TBC)
2. pneumococcus (pneumonia)
3. influenza (flu vaccination)

When did you receive a tuberculosis vaccination (BCG-TBC)?

When did you receive a pneumococcal vaccination?

When did you receive an influenza vaccination?

Do you smoke?
0. I have never smoked
1. on average, I smoke 20 cigarettes or more a day

2. on average, I smoke less than 20 cigarettes a day
3. I previously smoked an average of 20 cigarettes or more a day
4. I previously smoked an average of less than 20 cigarettes a day

Do you drink alcohol?

How often do you drink alcohol?
0. daily
1. several times a week
2. once a week
3. every two weeks
4. rarely (e.g. only at parties)

When you drink alcohol, how many alcoholic drinks do you drink?
0. 1-2 alcoholic drinks
1. 3-6 alcoholic drinks
2. 7 or more alcoholic drinks

Do you have pets?

What pets do you have?
0. fish/amphibians/reptiles
1. dog
2. rabbit
3. cat
4. guinea pig
5. bird
6. other (free text)

Has any pet shown 'signs of a cold'?

Are you interested in participating in our ELISA study and would you be willing and able to come to our study center on at least four study dates? Here, randomly selected participants will be screened for both current infection with the new coronavirus (throat swab/"PCR") and past infection and body defense response to the virus (blood draw/"antibody test"). You will be informed by e-mail within the next few weeks whether you have been selected.
0. I am interested and able to participate in the described examinations. I agree to be contacted for this purpose via the e-mail address used during registration
1. I am not interested or am not able to participate in the described investigations

**Follow-up questionnaire for ELISA participants**

Which of the following symptoms do you currently have?
0. none
1. fever / increased temperature >37.5 °C
2. cough
3. shortness of breath
4. muscle / limb pain
5. sore throat
6. cold
7. new onset of loss or change of smell
8. new onset of loss or change of taste
9. headache
10. nausea
11. vomiting
12. diarrhea
13. increased sweating
14. chills
15. seizures
16. fatigue/increased tiredness
17. dizziness
18. unsteadiness of stance and/or gait
19. other (free text)

Have you taken your temperature?

Please specify the measured body temperature

Where did you take your temperature?
0. in the mouth
1. in the armpit
2. in the anus
3. in the ear
4. in the groin

Do you have any sputum?

What does the sputum look like?
0. colorless/clear
1. whitish/mucous
2. yellowish/greenish
3. sanguineous
4. other (free text)

When do you experience shortness of breath?

0. during exertion
1. at rest
2. while lying down

Is your nose blocked?

Do you have a runny nose?

What does the nasal discharge look like?
0. colorless/clear
1. whitish/mucous
2. yellowish/greenish
3. sanguineous
4. other (free text)

In the last 72 hours, how often did you vomit?

During the last 72 hours, how often did you have bowel movements?

You have indicated that you currently have other symptoms. What are they?

Do you currently suffer from allergy symptoms (watery eyes, runny nose)?

In the last 72 hours, have you been in contact with one or more COVID-19 patient(s) who tested positive for COVID-19?

In the last 72 hours, have you been in contact with any person suffering from flu symptoms (e.g., fever, cough, or sore throat)?

Have you been tested for Corona virus infection in the last 72 hours?

Was the testing result positive?

Have you spent time in public areas in the last 72 hours?

Have you worn a mouth/nose mask in public in the last 72 hours?

Do you use an app that informs you about possible infection risk in connection with COVID-19?

Do any children under the age of 18 years live in your household?

Have any of these children been to school in the last 72 hours?
0. in daycare/kindergarten?
1. in elementary school?

2. in secondary school?
3. none

In the last 72 hours, did you leave your house to go to work?

How many hours per day did you work away from home?

How many colleagues did you have close (less than 1.5m distance) and long contact (more than 15 minutes) with at work per day?
0. 0-2
1. 3-5
2. 5-10
3. more than 10

How many people (e.g., customers or patients) did you have close (less than 1.5m distance) and long (more than 15 minutes) contact with per working day?
0. 0-2
1. 3-5
2. 5-10
3. more than 10

How many people (e.g., customers or patients) did you have close (less than 1.5m distance) and short (less than 15 minutes) contact with per working day?
0. 0-2
1. 3-5
2. 5-10
3. more than 10

In the last 72 hours, have you been to/at…
0. a restaurant or cafe?
1. a pub or bar?
2. a discotheque or club?
3. a cinema, theater or concert?
4. a gym?
5. a doctor's or physiotherapist's office?
6. a hairdresser or beauty salon?
7. none of the above

In the last 72 hours, have you been shopping...?
0. at a supermarket?
1. at a farmer's market?
2. at a hardware store?
3. at a bakery?
4. at any other retail store?
5. none of the above

In the last 72 hours, have you been to an event with...
0. less than 10 people
1. 11 to 50 people
2. 51 to 100 people
3. more than 100 people
4. none of the above

Have you used public transportation in the last 72 hours?
0. train
1. bus
2. train
3. taxi
4. shared use of vehicles (e.g. "car sharing")
5. none of the above

Have you been away from Schleswig-Holstein in the last 72 hours?

Have you been to Hamburg in the last 72 hours?

Describe your current well-being on a scale from 1-10

How often have you been troubled by worries in the last 72 hours?

How would you rate the quality of your sleep in the last 72 hours?

How tired or exhausted did you feel in the last 72 hours?

How strongly di you feel that you may have symptoms of infection with the Corona virus in the last
72 hours?

How would you rate your social integration in the last 72 hours?

How often did you communicate via digital means (phone/video conferencing/chat) in the last 72 hours?

Scale (1-10): 1 = Not at all; 10 = Very much

#### Supplement – References


1. Homolka S, Paulowski L, Andres S, et al. Two Pandemics, One Challenge-Leveraging Molecular Test Capacity of Tuberculosis Laboratories for Rapid COVID-19 Case-Finding. Emerg Infect Dis 2020;26(11):2549–54.

2. Beetz C, Skrahina V, Förster TM, et al. Rapid Large-Scale COVID-19 Testing During Shortages. Diagnostics (Basel) 2020;10(7):464.

3. World Health Organization. WHO Emergency Use Listing for In vitro diagnostics (IVDs) Detecting SARS-CoV-2 Nucleic Acid. https://www.who.int/diagnostics_laboratory/200908_eul_sars_cov2_product_list.pdf, assessed May 9, 2021.
